# Widespread adoption of precision anticancer therapies after implementation of pathologist-directed comprehensive genomic profiling across a large US health system

**DOI:** 10.1101/2024.01.02.23300311

**Authors:** Alexa K. Dowdell, Ryan Meng, Ann Vita, Bela Bapat, Douglas Hanes, Shu-Ching Chang, Lauren Harold, Mark Schmidt, Cliff Wong, Hoifung Poon, Brock Schroeder, Roshanthi Weerasinghe, Rom Leidner, Walter J. Urba, Carlo B. Bifulco, Brian D. Piening

## Abstract

Precision therapies and immunotherapies have revolutionized cancer care, resulting in significant gains in patient survival across tumor types. Despite this transformation in care, there is variability in the utilization of tumor molecular profiling. To standardize testing, we designed a pathologist-directed test ordering system at time of diagnosis utilizing a 523-gene DNA/RNA hybrid comprehensive genomic profiling (CGP) panel. We assessed actionability rates, therapy choices, and outcomes among 3,216 patients. 49% of cases had at least one actionable genomic biomarker-driven (GBD) approved and/or guideline-recommended targeted or immunotherapy and 53% of patients would have been eligible for a precision therapy clinical trial from three large basket trials. When assessing CGP versus an *insilico* 50 gene panel, 67% of tumors compared to 33% harbored actionable alterations. Among patients with 6-months or more of follow-up, over 52% received a targeted therapy or immunotherapy, versus 32% that received conventional chemotherapy alone, a phenomenon not previously observed.

**Statement of Significance:** This study represents the first report where precision therapies (targeted and immunotherapy) have overtaken traditional cytotoxic treatments in the routine community-based care of advanced cancer patients, resulting in better overall survival. This represents an important milestone in the evolution and adoption of precision oncology and highlights the importance of CGP.

## Introduction

Genomic biomarker-driven (GBD) personalized treatment regimens represents an important evolution in clinical cancer treatment over the last decade or more. Mutation-targeting therapies (*e.g.* BRAF/MEK inhibitors, EGFR tyrosine kinase inhibitors, TRK kinase inhibitors, poly-ADP ribose polymerase inhibitors (PARPi) etc.) have been associated with significant improvement in outcomes for patients with tumors harboring specific gene alterations^1–8^. In parallel, immune checkpoint inhibitor (ICI)-based immunotherapies have emerged as a major treatment modality that has resulted in remarkably durable responses in a subset of treated patients^9–20^. Of note, key biomarkers for ICIs include PD-L1 expression by immunohistochemistry, which is associated with improved response rates^21–24^; similar associations have been seen using genome-guided biomarkers such as tumor mutational burden (TMB) and microsatellite instability (MSI) assessment^21,22,25–29^. Notably, for each of these precision modalities, tumor genomic profiling is a key component to identify specific pathogenic mutations that can be targeted with these drugs. The field continues to evolve rapidly and now highly common driver genes, such as *KRAS,* are targetable with both precision and immunotherapeutic approaches^30–32^.

Given the rapidly evolving landscape of biomarker-guided therapies, a key challenge is enabling universal access to genomic testing that comprehensively covers all currently relevant genomic biomarkers. Key barriers to CGP access include the preponderance of labs that have not yet transitioned from older tumor profiling tools such as single-gene or small panel testing, cost of CGP, variable coverage of CGP by insurers, and physician hesitancy, possibly at least in part triggered by the complexity of the interpretation of CGP reports^33–35^. Despite these surmountable obstacles, adoption of CGP by laboratories around the world is rapidly increasing, with a variety of studies showing a clear benefit in identification of a broader spectrum of actionable targets and survival gains for precision medicine-treated patient populations^36–43^ Additional benefits realized by CGP testing include improved turnaround time and proper diagnostic tissue management and preservation, enabled by the assessment of all actionable markers in a single assay. CGP used as a pathologist-directed test post diagnosis at Providence also has the potential to aid the diagnostic process, as landmark tumor alterations or mutational signatures may resolve a differential diagnosis or add evidence to assign a tissue origin to a carcinoma of unknown primary (CUP). Moreover, we found inherent value in making tumor genomic profiles available to researchers to aid in the development of the next generation of precision oncological approaches. This is clear in the profound impact that The Cancer Genome Atlas (TCGA) has had on the field, and by innovations such as the more recent AACR-led community partnership Project GENIE (Genomics Evidence Neoplasia Information Exchange), which has already aggregated over 100,000 tumor genomics cases and is rapidly growing in scale^44–47^.

In order to systematically assess the impact of pathologist-directed CGP testing, we developed a novel programmatic approach where testing was initiated at time of diagnosis and performed at no cost to patients over a multi-year period. From this, we evaluated the actionability of the CGP results and analyzed the systemic therapies and outcomes of the CGP-tested patient cohort.

## Methods

### Study design and patient population

For the current study, we introduced two novel changes to standard clinical workflows at Providence. First, we instituted a pathologist-directed protocol system-wide, where somatic testing was performed immediately at time of diagnosis for all advanced solid tumor patients, and second, we standardized all testing to a single 523-gene DNA/RNA comprehensive genomic profiling (CGP) assay. Pathologist-directed CGP testing was instituted with the intent to reduce time between diagnosis and availability of CGP results, to inform clinical decision making, to standardize the testing methodology to a CAP CLIA certified platform, and to ensure the appropriate inclusion of all patients. This analysis included all patients tested with CGP under the pathologist-directed protocol in the Providence Molecular Genomics Laboratory between September 2019 and November 2021.

### IRB approval

All research was performed under protocol 201900048 “Effect of Automatic Reflex Genomics Testing on Clinical and Economic Outcomes in Cancer” approved by Providence IRB. All CGP results and associated clinical metadata were deidentified and aggregated for these analyses.

### Data availability statement

Genomic data analyzed in this study are available through cBioPortal from AACR Project GENIE under submitting institution Providence.

### Clinico-genomic dataset construction

We created an integrated dataset with curated and standardized fields. Biomarker data included variant call files (vcfs), annotations, designations of pathogenicity by pathologists/geneticists, clinical interpretations, sequencing quality metrics, and PD-L1 IHC results. Biomarker results were linked to clinical data extracted from both structured and unstructured electronic health records (EHR [Epic CareEverywhere]). Some data points (e.g., histology, treatment status) were curated manually. Statistics were conducted on the aggregate patient cohort to determine the median and interquartile range (IQR) for patient age, test turnaround time, and follow up time since the test was ordered.

### Data extraction and utilization of Snowflake

Pooled electronic medical records and genomic data were curated, standardized, and stored on a cloud data warehouse. Patients who received testing between September 2019 and November 2021 were selected for further analysis (N = 3,216 patients). Of those 3,216 patients, 2,028 patients (63%) had reported follow-up or treatment information in our EMR, while the other patients likely sought treatment elsewhere, did not have EMR from their institution, or were lost to follow-up.

### Machine learning-based chart mining

To extract PD-L1 IHC results from clinical notes, we developed a machine learning-based information extraction system, which uses a customized version of spaCy^48^ for sentence segmentation and NLTK^49^ for tokenization. For key result extraction, our system uses domain-specific rules to identify relevant entities and potential cross-sentence relations, as well as to determine if they are positive assertions. Each extracted result contains the PD-L1 gene mentions, assay types (22C3, 28–8, SP263, SP142), staining intensity, analysis methods (tumor proportional score or combined positive score) and expression values (e.g., <1%, 10, high). We used the note date as a proxy for the measurement date, but we plan to extract the measurement date from the note text in future work. Our pipeline runs daily on new pathology reports, imaging reports, progress notes, encounter notes, op notes, and surgery notes, but we find that most PD-L1 results are reported in pathology reports and progress notes.

We developed our rules using notes prior to a certain date and constructed a test set from pathology reports and progress notes after that date. For the test set, we randomly selected 135 cancer patients and sampled one note per patient that contains a PD-L1 mention in any of its surface forms (e.g., PDL1, PDL-1, PDL 1). We asked an expert to manually annotate each sampled note for PD-L1 expression values and the analysis methods. We evaluated our pipeline by comparing the extracted expression values and methods with the expert annotations. The evaluation results show that our pipeline achieved high performance: precision 97.5, recall 89.6, and F1 93.4.

### Actionability assessment

Actionability was assessed based on criteria compiled in the OncoKB database^50^, a curated database that classifies all tumor-specific mutations into tiers of actionability. Briefly, tumor types were translated to OncoTree codes, assessed for stage, and matched to actionable biomarker categories (OncoKB levels 1 and 2). In order to characterize the subset of cases with potentially actionable markers for study, we compiled all reported pathogenic variants (e.g. SNVs, indels, fusions, CNVs) from the cohort and assessed potential actionability based on OncoKB therapeutic levels of evidence^50^. Patients were assigned to the level of most significant alteration based on levels of evidence: actionable biomarkers predictive of response to FDA approved therapies (level 1), or predict response to guideline recommended, or standard of care therapies (level 2). In instances where a biomarker became actionable within the window of our study (e.g. *KRAS* G12C in 2021), patients who received genetic testing before the FDA approval date of the specific treatment were not considered actionable, even if they harbored the actionable alteration.

To assess clinical trial matching eligibility, the patients were compared to the enrollment criteria for three basket trials (ASCO-TAPUR, NCI-MATCH, and MyPathway). If a patient met the genomic and cancer type enrollment criteria, and their date of testing fell within the trial’s eligibility timeframe, they were counted as eligible for the trial. Trials were not treated as mutually exclusive, and patients could be eligible for multiple clinical trials based on their genomic data and cancer type. The ASCO-TAPUR clinical trial arm for treatment with cetuximab in patients with wildtype *KRAS, NRAS*, and *BRAF* was left out of the analysis to prevent a scenario where all patients would be eligible for a clinical trial regardless of whether they possessed wildtype or mutated *KRAS*/*NRAS*.

A key putative benefit of CGP testing versus prior-generation small panels (e.g., 5-50 genes) is that more patients will be eligible for precision therapies due to the expanded biomarker spectrum in CGP. To test this, we assessed actionability in the cohort based on the original set of 523 genes in the CGP assay versus the subset of actionable mutations that would have been detected using a legacy 50-gene panel that was previously deployed at Providence. As such, we bioinformatically subset the CGP dataset to only the genes in the 50-gene panel and calculated the difference in actionable alterations between CGP and the *in silico* small panel.

### Treatment selection

We assessed therapy selection post CGP testing and subsequent clinical outcomes among patients with advanced/metastatic disease who received care at Providence. Treatments were classified by type of therapy: 1) matched targeted therapy (matched TT) defined as FDA approved and/or NCCN guideline based therapies requiring OncoKB level 1 and 2 biomarkers (e.g., Osimertinib for *EGFR*-positive lung cancer or pembrolizumab for TMB-high cancers); 2) Non-CGP biomarker associated TT (NBTT) are precision therapies that do not require actionable OncoKB level 1, 2 biomarkers (e.g., sorafenib in kidney cancer or everolimus in breast cancer); 3) Off-label TT are therapies that were administered to patients who lacked the matched OncoKB level 1 or 2 biomarker for the targeted therapy; 4) Immunotherapy (IO) including immune checkpoint inhibitors (ICI); 5) Chemotherapy and IO (chemo +IO) combination therapy of ICI and traditional anti-neoplastics; 6) Chemotherapy only includes use of traditional anti-neoplastics.

### Clinical outcomes

Overall survival (OS) was documented for all tumor types and separately for a subset of NSCLC patients. OS was classified as the duration in months from the date of report for the most recent CGP to the date of last follow-up or death from any cause. OS by treatment type is presented in unadjusted Kaplan-Meier figures and subsequently tested using a Cox proportional-hazards model adjusted for patient age and tumor type. All survival analyses were carried out in R v.4.2.2 using the survival package.

## Results

The goal of the study was to assess the impact of adoption of in-house CGP testing using a protocol where CGP testing was initiated by a pathologist immediately upon a histopathological advanced cancer diagnosis (Figure 1A). This workflow replaced a prior methodology largely utilizing small panel testing (5-50 genes), that is ordered by a medical oncologist at the time of therapy selection. By shifting NGS ordering upstream, we could facilitate 1) use of the NGS test results in the diagnostic process, and 2) make the expediting availability of NGS results as soon as possible to the treating physician. Testing was also performed under the research protocol at no cost to the patient, or the practicing oncologist’s clinic, to remove potential reimbursement-related barriers to CGP testing. A wide variety of tumor types were tested (Figure 1B), with the most common being lung (30%), bowel/colon (16%), breast (9%), pancreas (6%), and prostate cancer (5%). We observed a turnaround time of 14 days (IQR 12-15 days, Table 1), which was similar to prior small panel testing at the same institution. However, NGS results were available earlier in the clinical decision-making process; NGS results were available more frequently at the time of the initial medical oncologist visit (median −12 days) compared to prior non-pathologist-directed testing (median 0 days, p<0.005) (Figure 1C).

**Figure 1.**
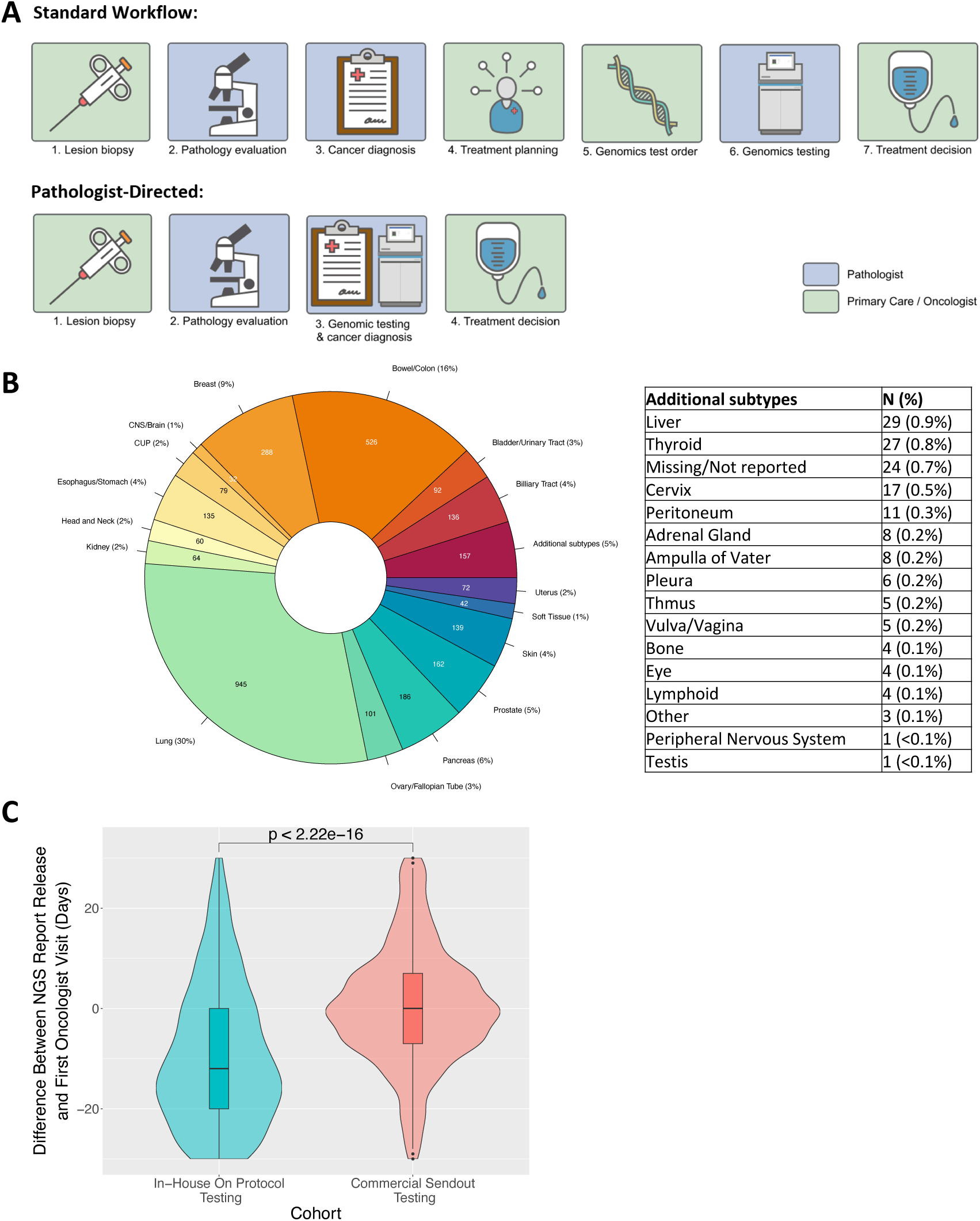
Overview of testing protocol and cancer subtypes. A) Pathologist-directed testing versus previous standard genomic testing workflows in oncology. B) Tumor type breakdown of the patients tested in this cohort. C) Time difference between NGS report release date and first oncologist visit, in days, between pathologist-directed testing (N=432 patients) and non-pathologist-directed testing (N=1,192 patients).

**Table 1.** Patient Characteristics.

|  | **All Patients** | **Stage IV Treated Patients** |
| --- | --- | --- |
| **Advanced cancer patients tested using CGP** | **3,216** | **1,482** |
| **Age at time of testing, median (IQR)** | 67.0 (60.0, 75.0) | 66.0 (58.0, 74.0) |
| **Sex, n (%)** |  |  |
| Female | 1,684 (52.4) | 779 (52.6) |
| Male | 1,532 (47.6) | 703 (47.4) |
| **Race, n (%)** |  |  |
| White | 2,592 (80.6) | 1,215 (82.0) |
| Black | 76 (2.4) | 33 (2.2) |
| Asian | 149 (4.6) | 87 (5.9) |
| American Indian or Alaska Native | 35 (1.1) | 15 (1.0) |
| Native Hawaiian or Other Pacific Islander | 21 (0.7) | 6 (0.4) |
| Unknown | 343 (10.7) | 126 (8.5) |
| **Stage, n (%)** |  |  |
| Stage III | 401 (12.5) | - (-) |
| Stage IV | 2,815 (87.5) | 1,482 (100.0) |
| **Year Test ordered, n (%)** |  |  |
| 2019 | 85 (2.6) | 45 (3.0) |
| 2020 | 1,359 (42.3) | 664 (44.8) |
| 2021 | 1,772 (55.1) | 773 (52.2) |
| **Turnaround time in days\*, median (IQR)** | 14.0 (12.0, 15.0) | 14.0 (12.0, 15.0) |
| **Follow-up in months since testing ordered\*\*, median (IQR)** | 11.0 (3.0, 19.0) | 15.0 (8.0, 21.0) |
| **Deceased, n (%)** | 1,508 (47.1) | 693 (46.8) |
| **Progression^, n (%)** | 495 (15.4) | 384 (25.9) |

Source: Providence Health System, 2019-2021
\*n=24 missing values were excluded
\*\*n=20 missing values were excluded
IQR= interquartile range; NSCLC= non-small cell lung cancer
^ Derived from MSFT HealthNext Progression Model, Cancer Registry Data and EMR treatment discontinuation documentation

The study included 3,216 patients with advanced solid cancer whose tumors were subject to CGP between September 2019 and November 2021 (Table 1). The median age of patients was 67 years, 52% were female, 80% were white. Most patients had metastatic disease (88%), 40% entered hospice, and 33% died during follow-up (median 5 months). Most of the 3,216 patients were tested in 2020 (42%) and 2021 (55%).

### Results of Actionability Assessment

Overall, 49% of CGP-tested cases had at least one guideline-based actionable biomarker (Figure 2A). Figure 2B shows the breakdown of mutations identified for genes that are actionable based on FDA or standard of care biomarkers (OncoKB 1 and 2). We observed that high tumor mutational burden (TMB-High) was the single actionable biomarker with the highest number of cases (22%) and was detected across almost all the main tumor types tested, followed by BRAF (11%), and EGFR (4%). Consistent with the literature, we also observed subsets of pan-cancer biomarkers enriched across a broad set of tumor types (e.g. microsatellite instability high (MSI-High) and *NTRK1-3* fusions) as well as biomarkers that were highly specific to a subset of tumor types (e.g. *BRAF* in melanoma and colorectal cancer and *BRCA1/2* in ovarian cancer). Figure 2C shows the distribution of all detected pathogenic alterations (i.e., not filtered by current actionability). As expected, we observed high mutational rates for common tumor suppressor genes such as TP53. Additionally, we identified a long tail of other clinically significant genes that may be targetable in the future.

**Figure 2.**
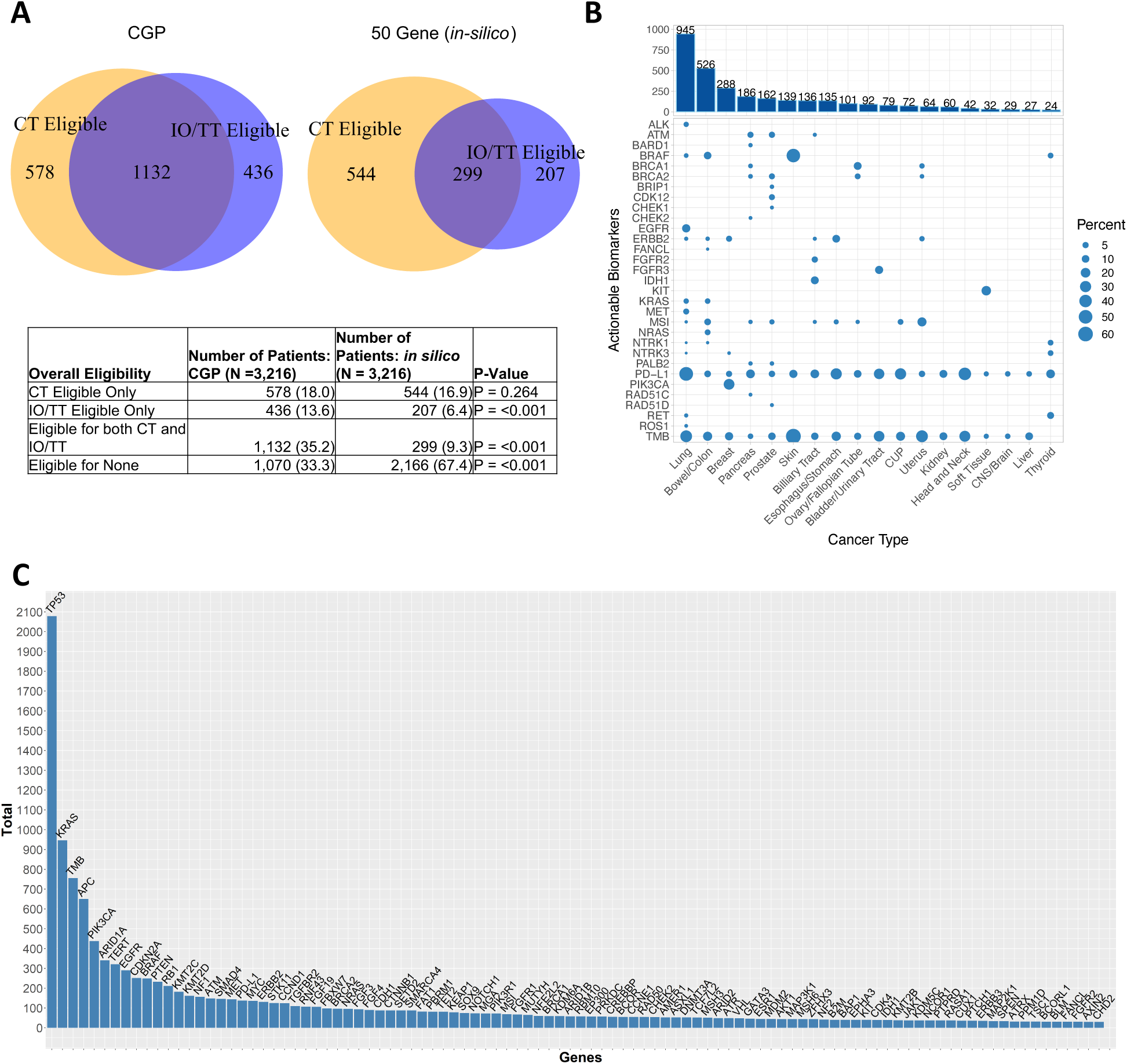
Mutational actionability/pathogenicity in the cohort. A) Comparison of the proportion of guidelines-based IO/TT or clinical trial-based IO/TT eligibility for CGP tested patients versus 50-gene *in silico* subset panels. B) Frequency map showing the number of cases per tumor type with actionable mutations in the top evidence categories (OncoKB levels 1/2/R1). C) Long-tail histogram plot for the top 100 pathogenic mutations across all cases.

Overall, 53% of patients tested with CGP matched to one or more arms across the three assessed basket clinical trials (ASCO-TAPUR, NCI-MATCH, and MyPathway), compared to 26% (p<0.001) if patients were tested using only a 50-gene panel (*in silico* cohort). Matched cases included patients matching to immunotherapy trial arms based on TMB-High but also cases matching based on rare biomarkers (*NTRK*, *PTCH1* etc.). Overall, 67% of tumors from CGP-tested cases compared to 33% for *insilico* 50 gene panel cases (p<0.001) harbored actionable mutations based on either guideline-based or clinical trial matching.

### Treatment utilization across CGP-tested cases

While the prior analyses determined which cases were potentially actionable based on genomic biomarkers, precision and IO therapies represent just a subset of treatment modalities available to the treating oncologist and patient. As such, we next assessed the frequency of utilization of precision/IO therapies for cases with actionable mutations as compared to traditional anti-cancer therapies (*e.g.,* chemotherapy). From the original 3,216 patients in the cohort, 1,482 patients with documented clinical follow up and treatment in our EMR were evaluated for treatment selection and outcome. Figure 3 shows treatment type utilization across the cohort. Of note, precision/IO therapy utilization was high, with 54% of patients with an appropriate IO biomarker receiving either IO or IO plus chemotherapy. This far exceeded the percentage of tested patients that received chemotherapy alone (32%). We also grouped treated patients based on what types of biomarkers were detected in their CGP profile (Figure 3). In patients with an actionable targeted therapy (TT) biomarker, use of a TT was indeed the most widely used single modality, with 59% of TT-positive patients receiving a targeted therapy. Similarly, for patients with IO-positive biomarkers (TMB-High, MSI-High, or PD-L1 positive), IO was the dominant treatment modality, with 63% of patients receiving an immunotherapy with or without chemotherapy. As expected, we detected little targeted therapy use in patients with no actionable mutations in their CGP profile (33 patients, 2.0% of cohort). Off-label TT use was even more rare in cases without an actionable TT biomarker or a positive IO biomarker (15 cases 1% of cohort), suggesting that off-label use of a TT may have occurred in rare cases where all other treatment options were exhausted. Immunotherapy use in cases without an IO-positive biomarker was more common, with 13% receiving IO without a matched biomarker, indicative of newer IO treatment guidelines in advanced cancer without biomarker indications.

**Figure 3.**
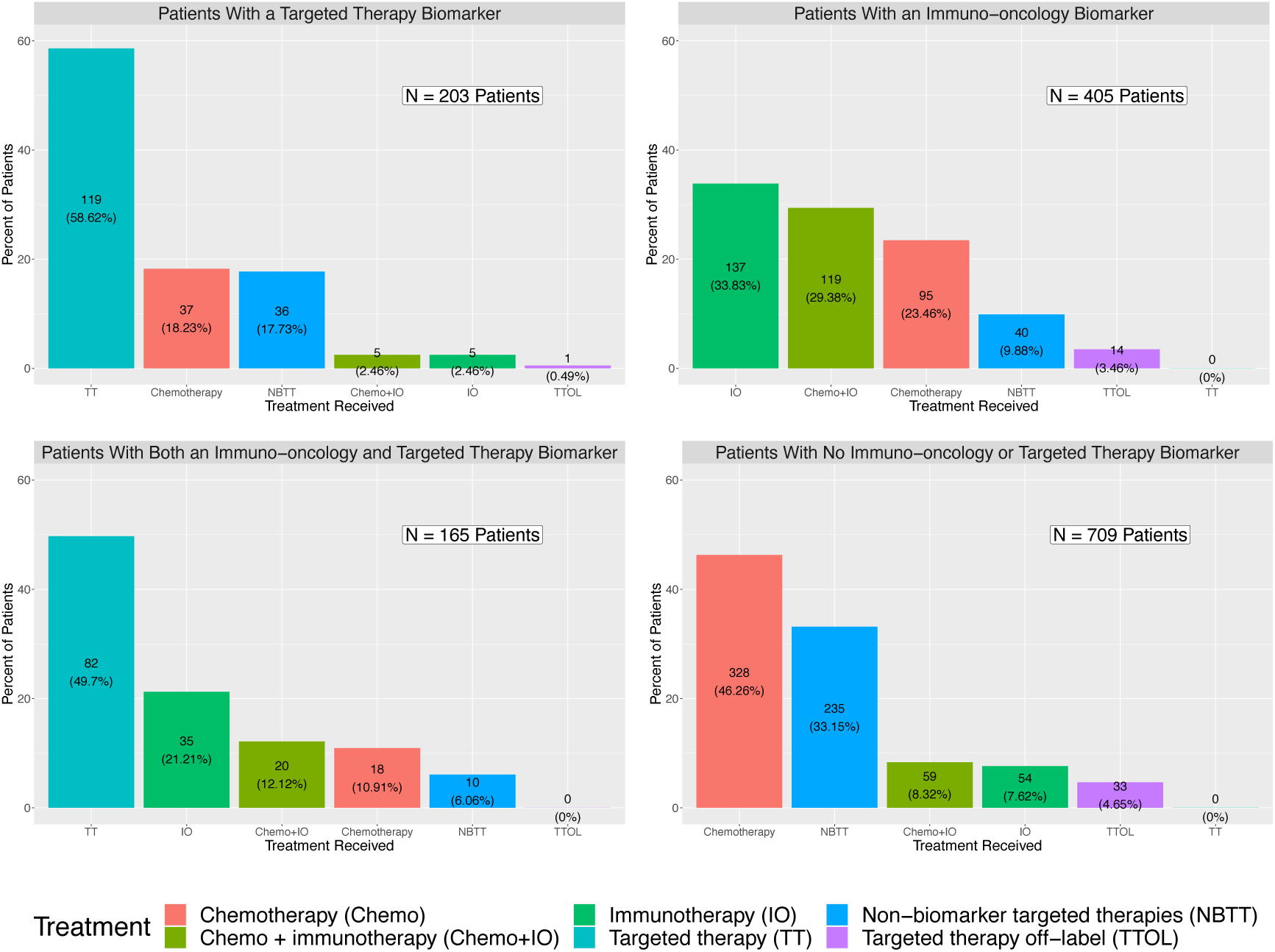
Distribution of types of therapy post-testing. Chemotherapy (Chemo), immunotherapy (IO), chemo+IO, targeted therapy (TT), non-biomarker targeted therapies (NBTT), and targeted therapy off-label (TTOL) use for patients with documented follow up, grouped by the presence or absence of TT or IO biomarkers.

### Clinical outcomes for CGP-tested patients

Across all tumor types, patients treated with biomarker-guided TT or IO show significant improvements in overall survival versus chemotherapy (Figure 4A&B). Median overall survival was 25 months (targeted therapy) vs. 17 months (chemotherapy only). Patients receiving targeted therapy had better survival outcomes compared to chemotherapy only (HR 0.66, p<0.001), even after controlling for age and tumor type. For the subset of patients diagnosed with NSCLC, the median overall survival was 26 months (targeted therapy) vs. 16 months (chemotherapy only). NSCLC patients receiving targeted therapy had better survival outcomes compared to chemotherapy only (HR 0.55, p=0.032) after controlling for age. While the survival benefit of targeted therapies has been shown previously in a variety of contexts, this represents the first large-scale evaluation in a community setting with an in-house CGP testing program.

**Figure 4.**
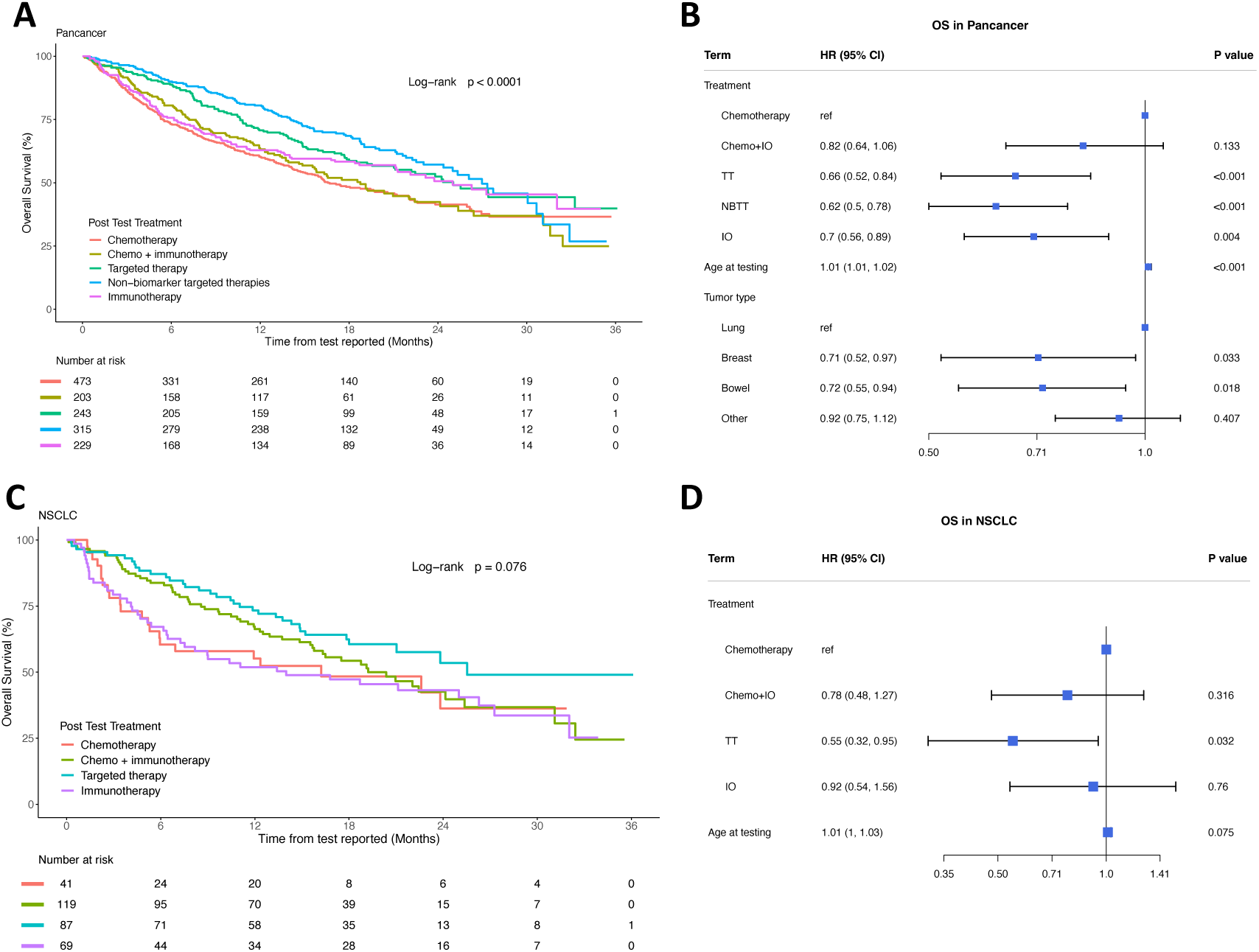
Overall survival in the CGP cohort. A) Pan-cancer Kaplan-Meier curve showing survival post CGP-testing for patients treated with different modalities. B) Pan-cancer hazard model for treated patients. C) Non-small cell lung cancer (NSCLC) Kaplan-Meier survival curves for different treatment modalities. D) NSCLC hazard model for treated patients.

## Discussion

Our study represents a unique look at the impact of a programmatic deployment of in-house, pathologist-directed CGP testing across a community health system with diverse clinical practice. Given the widespread diversity in genomic testing utilization in the community setting, we proposed that this standardization to a single comprehensive genomic panel may yield significant benefits in uncovering new targeted therapy options for patients that may otherwise receive conventional chemotherapy. Indeed, we show that among CGP-tested patients, 49% had one or more actionable biomarkers for a targeted therapy or immunotherapy, a result that significantly exceeded the actionable fraction when we retrospectively evaluated the same patients *in silico* using a legacy 50-gene panel, suggesting that a significant subset of patients receiving small panel testing would likely benefit from being retested with a CGP. Participation in the CGP program at Providence was voluntary for individual clinics and physicians, and while a significant proportion of the health systems did participate, those that did not suggest the existence of additional barriers to adoption of CGP testing as a major modality. A key follow-up health improvement study will be to engage with the physician community to identify the key factors driving reluctance to adopt CGP in their practices. Additionally, CGP testing was provided in a context that introduced no new charges for the patient, but in a real-world scenario, payers would need to subsidize CGP testing to improve overall population outcomes and to accelerate utilization of CGP in a clinical setting.

A key finding among the CGP-tested cohort was that 67% of patients, regardless of biomarker status, received some type of targeted therapy or immunotherapy. While some of these were guideline based and not linked to a specific biomarker (e.g., VEGF inhibitors, CDK inhibitors etc.), the majority were biomarker-driven. Regardless of indication, the widespread use of targeted modalities versus chemotherapy alone represents an important paradigm shift in the treatment of advanced cancer and is a testament to the increasingly wide spectrum of targetable oncogenic alterations and immunological modulators that have made oncology the standard-bearer for precision approaches in medicine.

Despite these successes, there are still significant gaps in providing precision medicine for all advanced cancer patients. A significant proportion of CGP results still do not yield an actionable marker, and that number is particularly high across a subset of tumor types. While some key frequent targets have recently become actionable (*KRAS, IDH1* etc.), clearly more research and development are needed for precision anti-cancer molecular therapies for the still significant subset of tumors with non-actionable genomic findings. Among patients treated with targeted therapies, more work is needed in improving efficacy of these drugs, developing alternative therapies to address acquired resistance, and developing effective multi-target combinations to potentially preclude tumor evolutionary escape. For the subset of patients with actionable markers that did not receive a precision therapy, significant effort needs to be invested in education of physicians and in facilitating access to treatment. Given the high cost of many of these treatments, and the substantial staffing necessary to assure prior authorization, socioeconomically disadvantaged communities may be particularly susceptible to lack of access. As such, novel programs and strategies are needed to ensure that precision medicine is truly available to all who need it.

Of note, what we call “comprehensive” today in terms of CGP testing likely will be supplanted by even more extensive panels or somatic whole genome sequencing as more actionable targets emerge and sequencing costs continue to decrease. While this is predicted to be associated with increased benefit through access to precision therapies, and decreased conventional toxicity associated with chemotherapy, additional complexities arise in terms of potentially choosing among multiple independent precision options for a single patient (a phenomenon that is becoming increasingly common today). As such additional clinical trial work is underway evaluating sequencing of multiple therapies and/or combinations of these therapies (i.e. NCI ComboMATCH). Given the rapid increase in precision medicine developments coupled with rapidly decreasing sequencing costs, we expect this era is not far away.

## Acknowledgements

This work is dedicated to the memory of our friend and colleague Mark Schmidt. We thank members of the Molecular Genomics Laboratory at Providence for performing all molecular testing associated with this study as well as pathologists across Providence for their participation in the study. B.B. and B.S. are employees of Illumina, Inc; San Diego, CA. C.W. and H.P. are employees of Microsoft Corp; Redmond, WA. The study was funded in part by Illumina.

